# Experience from a COVID-19 screening centre of a tertiary care institution: A retrospective hospital-based study

**DOI:** 10.1101/2020.11.01.20224105

**Authors:** Somen Kumar Pradhan, Dinesh Prasad Sahu, Durgesh Prasad Sahoo, Arvind Kumar Singh, Binod Kumar Patro, Sachidananda Mohanty

**Affiliations:** Department of Community Medicine and Family Medicine, All India Institute of Medical Sciences, Bhubaneswar, Odisha, India; Medical Superintendent, All India Institute of Medical Sciences, Bhubaneswar, Odisha, India

**Author notes:** **Corresponding author:** Binod Kumar Patro, Address: Third Floor, Academic block, All India Institute of Medical Sciences, Bhubaneswar. Odisha, India-751019. Contact Number: 9438884013. Email-Id.

**Keywords:** COVID-19, Screening OPD, Tertiary care hospital

## Abstract

**Introduction:** The COVID-19 pandemic continuing to be a significant public health concern across the whole world, including India. In the absence of any specific treatment or vaccine against COVID-19., the role of efficient testing and reporting has been uncontested so far as the number of cases is rising daily. In order to strengthen the screening activities and to prevent nosocomial infection, facility-based screening centres have been designed and operated at various level of healthcare, including tertiary care institutions.

**Methods:** The present study has been planned with an objective to understand the patient profile and evaluate the functioning of COVID-19 screening OPD(CS-OPD) at a tertiary care hospital. In this hospital-based retrospective study, data from individuals visiting the COVID-19 screening OPD during the period from 17th March 2020 to 31st July 2020 were collected. We documented and analysed relevant demographic, epidemiological and clinical characteristics of the patients.

**Results:** A total of 10,735 patients visited the COVID-19 screening OPD during the defined study period out of which 3652 individuals were tested. Majority of the patients, i.e. 65.67% (7050) were male and in 15-59 years age group (84.68%). Most common symptoms among patients visiting CS-OPD was Cough (9.86%). Out of the total, 17.17% (1843) of patient reported to the CS-OPD with contact history of COVID-19 positive patient. On the other hand, 13.49% (1448) of patients were with either domestic or international travel history. The overall testing rate and positivity rate for CS-OPD during this period were found to be 34.02% and 7.94% respectively.

**Conclusion:** The clinical, demographic and epidemiological characteristics of patients visiting CS-OPD varied across the study period depending upon the containment and testing strategy. The CS-OPD played a crucial role in preventing nosocomial infection and maintaining non-COVID care at the tertiary care hospital.

## Background

The novel severe acute respiratory syndrome coronavirus 2 (SARS-CoV-2, previously known as 2019-nCoV, also known as COVID-19) has rapidly spread in India, with unprecedented propagation, because of its highly infectious nature. The World Health Organization declared the COVID-19 outbreak as a pandemic on 11^th^ March 2020. On 30^th^ January 2020, the first confirmed case of COVID-19 was detected in India. (1) The state of Odisha identified the first case of COVID-19 on 15^th^ March 2020. (2) With a rising number of cases, the need to screen all patients with respiratory symptoms and travel history was recognised. One effective strategy was the establishment of fever clinics or COVID-19 screening centres. These screening centres were assigned to screen patients based on a standard criterion. With increasing instances of nosocomial outbreaks of COVID-19, it has become even more important to screen all patients with suspected infectious disease in the hospital setting and for control and prevention of infection in the community. (3)

As pandemic alert was sounded in India and patient screening in health facilities was strongly recommended, Ministry of Health and Family Welfare (MOHFW), India issued guidelines for setting up COVID-19 screening centres at healthcare settings. The purpose of these Screening Centre was to (a) Attend to patients of influenza-like illness in a separate area from the general outpatient department(OPD),(b) To facilitate implementing standard droplet precautions,(c) To triage the patients,(d) Collect samples. (4)

Based on these principles, health care institutions have developed and implemented a hospital-specific systematic process for screening and managing suspected COVID-19 patients. (5)(6) However, till now there is limited published literature regarding functioning and patient profile of these COVID-19 screening facilities, especially at tertiary care institutions which are significantly involved in both COVID-19 and non-COVID-19 services simultaneously. (7) Hence, we planned this study to understand the patient profile and evaluation of COVID-19 screening centres at a tertiary health care institution.

## Materials and methods

This study was conducted in the COVID-19 screening Outpatient department (CS-OPD) under the Department of Community & Family Medicine at All India Institute of Medical Sciences (AIIMS), Bhubaneswar. AIIMS Bhubaneswar is a 922 bedded tertiary care institution in the state of Odisha which caters for approximately 1.2 million outpatients, 30,000 inpatients and 40,000 emergencies per year.

The AIIMS, Bhubaneswar CS-OPD was made functional from 17^th^ March 2020. The main objective of CS-OPD was to maintain the function of the tertiary care institution through segregating COVID-19 and Non-COVID-19 patients through screening before their admission or entry into the hospital. The functioning of the CS-OPD was based on standard operating procedures devised by the COVID-19 management committee of the institution with the help of all the guidelines and protocol notified by Indian Council of medical research (ICMR) and MOHFW. A two-level of screening system was made operational as the number of cases started to rise in the state. As part of the first level of screening, all the patients and their attendants were screened at the entry gate of main OPD by paramedical staffs based on three criteria, i.e. travel history, contact history and complaint of fever (by measuring body temperature with the help of infrared thermometer). Patients needing immediate hospitalisation were referred to Trauma and Emergency ward for screening and treatment. Based on the first level triage result, the patient was either referred to CS-OPD for second-level screening by medical officers (if any of the suspect criteria was present) or inside the main OPD building for further treatment (if none of the suspect criteria was present). Patient or visitor could also directly visit the CS-OPD and was also referred by clinicians from their OPD. From 15^th^ June 2020 onwards, all the patients requiring admission, surgical interventions or day-care procedures were also referred to CS-OPD for COVID-19 testing.

The CS-OPD was situated in a standalone building in the peripheral zone of hospital separated from other general OPDs with separate entry and exit point to prevent possible cross-infection. The CS-OPD was broadly divided into the following four zones: Registration-cum-report collection zone, OPD zone, Sample collection zone and Control room-cum-nursing station zone. **(Figure-1)** A total of 21 staffs from the existing pool of AIIMS, Bhubaneswar has been designated to work at CS-OPD and to ensure its proper functioning. **(Table-1)**. Patient at CS-OPD/she was managed as per the algorithm explained in Figure-2. **(Figure-2)** The CS-OPD was operational from 8.00 AM to 2.00 PM every day. Patients were categorised into “Suspect” and “Not-a-suspect” case for COVID-19 based on travel history (In last 14 days), contact history (In last 14 days) and relevant symptoms suggestive of COVID-19 (fever, sore throat, cough, dyspnoea, loss of taste/smell). A suspected case was categorised into ICMR specified categories and referred to the sample collection zone. **(Figure-3)** The nasopharyngeal swab samples are collected in two sessions, i.e. 8.00 AM-11.00 AM and 11.00 AM-2.00 PM. All the samples are sent to the ICMR approved Viral Research and Diagnostic Laboratory (VRDL) of microbiology department under a proper cold-chain system. All the samples are tested by Reverse Transcriptase-Polymerase chain reaction (RT-PCR) method.

**Figure 1:**
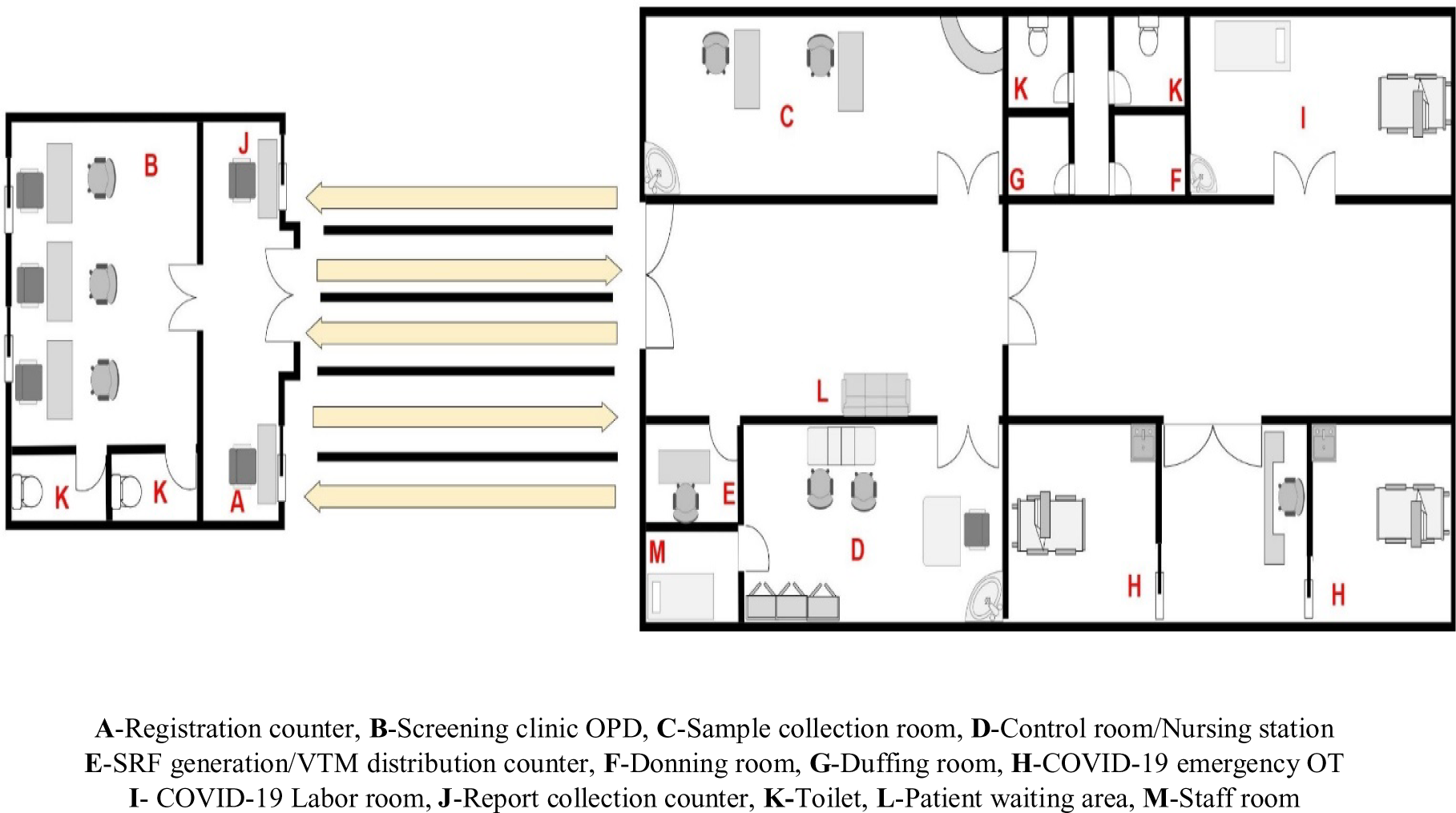
COVID-19 screening OPD (CS-OPD) map:

**Table 1:** Human resources for CS-OPD-.

| Sl.No. | Designation | Number |
| --- | --- | --- |
| 1 | Faculty-in-charge | 1 |
| 2 | Senior Resident (Doctor) | 2 |
| 3 | Junior Resident (Doctor) | 2 |
| 4 | Intern | 2 |
| 5 | Nursing officer | 6 |
| 6 | Lab technician | 1 |
| 7 | Data entry operator (DEO) | 2 |
| 8 | Medical social worker (MSW) | 2 |
| 9 | Security staff | 2 |
| 10 | Housekeeping staff | 1 |

**Figure 2:**
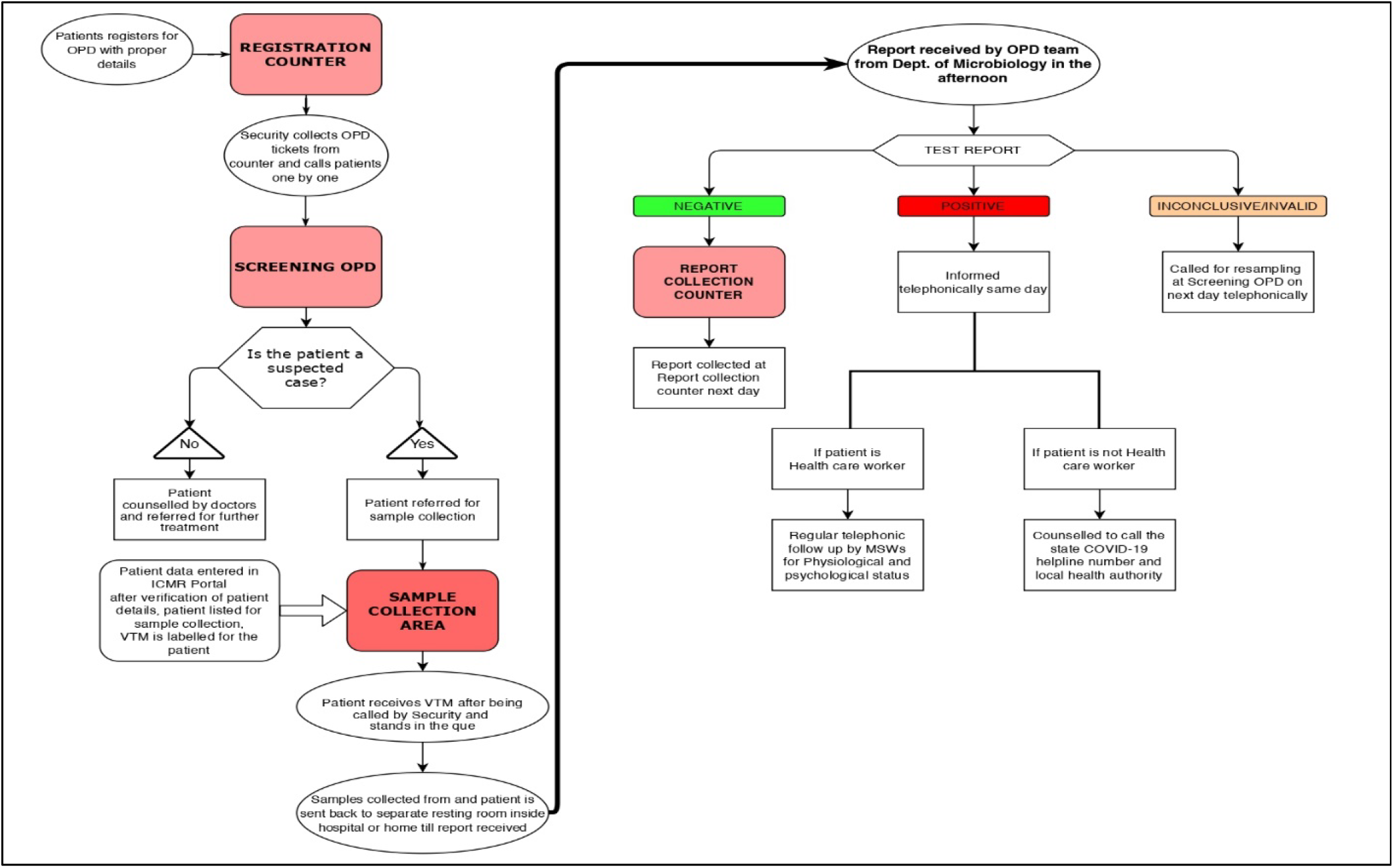
Algorithm for patient-flow and service delivery at CS-OPD.

The present study is a hospital-based retrospective study in which all the patient-related data of subjects attending COVID-19 screening OPD during 17^th^ March 2020 to 31^st^ July 2020 were collected and analysed. Any patient with incomplete or missing data or duplicity were being excluded from the study. At the time of patient examination, the staffs input the data, such as the presence of COVID-19 symptoms, travel history and a history of contact with COVID-19 patients, after interviewing the patient. The study used standard case definitions and categories for sample collection as advised by the MOHFW and ICMR. (8)(9) **(Figure-3)**

**Figure 3:**
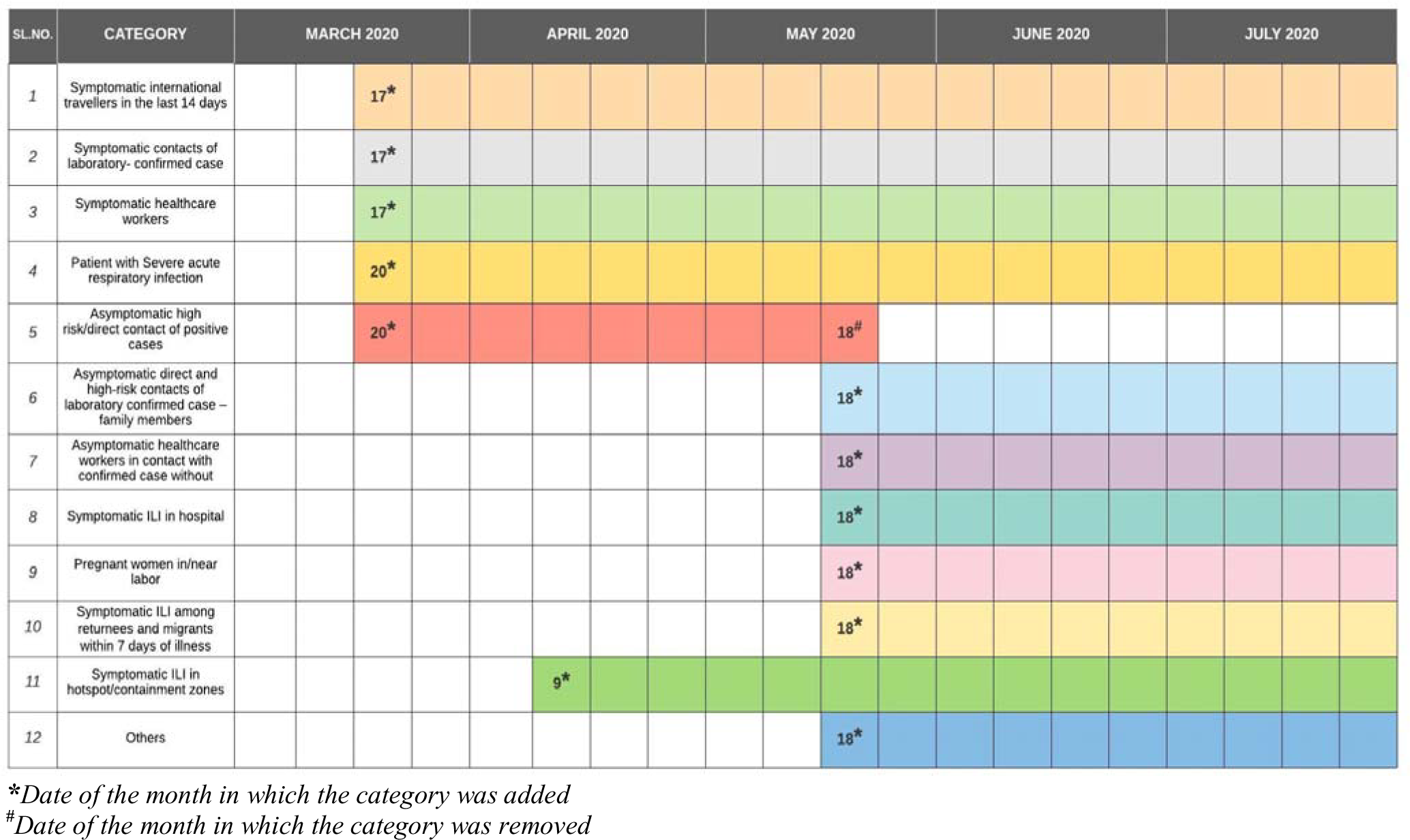
Timeline of revision of COVID-19 testing strategy by ICMR:

All the extracted quantitative data were administered in Microsoft excel along with the relevant variables mentioned. Data analysis was performed by SPSS version 23. Analysis of socio-demographic variables as well as variables related to COVID-19 screening services was expressed using descriptive statistics like mean, median, proportion and relevant graphical presentation. Personal identifiers for the patients were removed from the data set after data extraction to maintain privacy and confidentiality. The institutional ethical committee of AIIMS, Bhubaneswar approved this study (Reference number: T/IM-NF/CM&FM/20/72).

## Results

A total of 10,735 patients were screened for COVID-19 during these 136 days (17^th^ March 2020-31^st^ July 2020). A median of 57 (IQR=93.00) patients visited CS-OPD every day. Out of them, majority, i.e. 65.67% (7050) were male and in 15-59 years age group (84.68%). The median age among male and female patients visiting CS-OPD was 34.00 (IQR=22.00) and (IQR=20.00) years, respectively. Most common symptoms among patients visiting CS-OPD was Cough (9.86%) followed by fever (9.30%). However,79.23% (8505) of the patients were without any specific symptoms relevant to COVID-19. Out of all, 17.17% (1843) patient reported to the CS-OPD with contact history of COVID-19 positive patient. 13.49% (1448) of patients were with either domestic or international travel history within the last 14 days of visit to CS-OPD. **(Table-2)**

The seven days moving average for the number of patients with travel history was higher initially in March 2020 (range:2.4-8.4) followed by a decrease during April 2020(range:0.7-1.7). However, it showed an increasing trend from May 2020 till the first week of July 2020 (range:1.7-42.1). Similarly, the seven days moving average for the number of patients with contact history was stagnant during March 2020 to May 2020 (range:0-4.6). But it peaked during June 2020 and the first week of July (range:1.6-73.3). Similarly, for the number of patients with symptoms was initially higher in March 2020 (range:12-28) followed by a decline from April 2020 to May 2020 (range:6-14). But It further increased during June 2020 to July 2020 (range: 3-37) **(Figure-4)**

**Figure 4:**
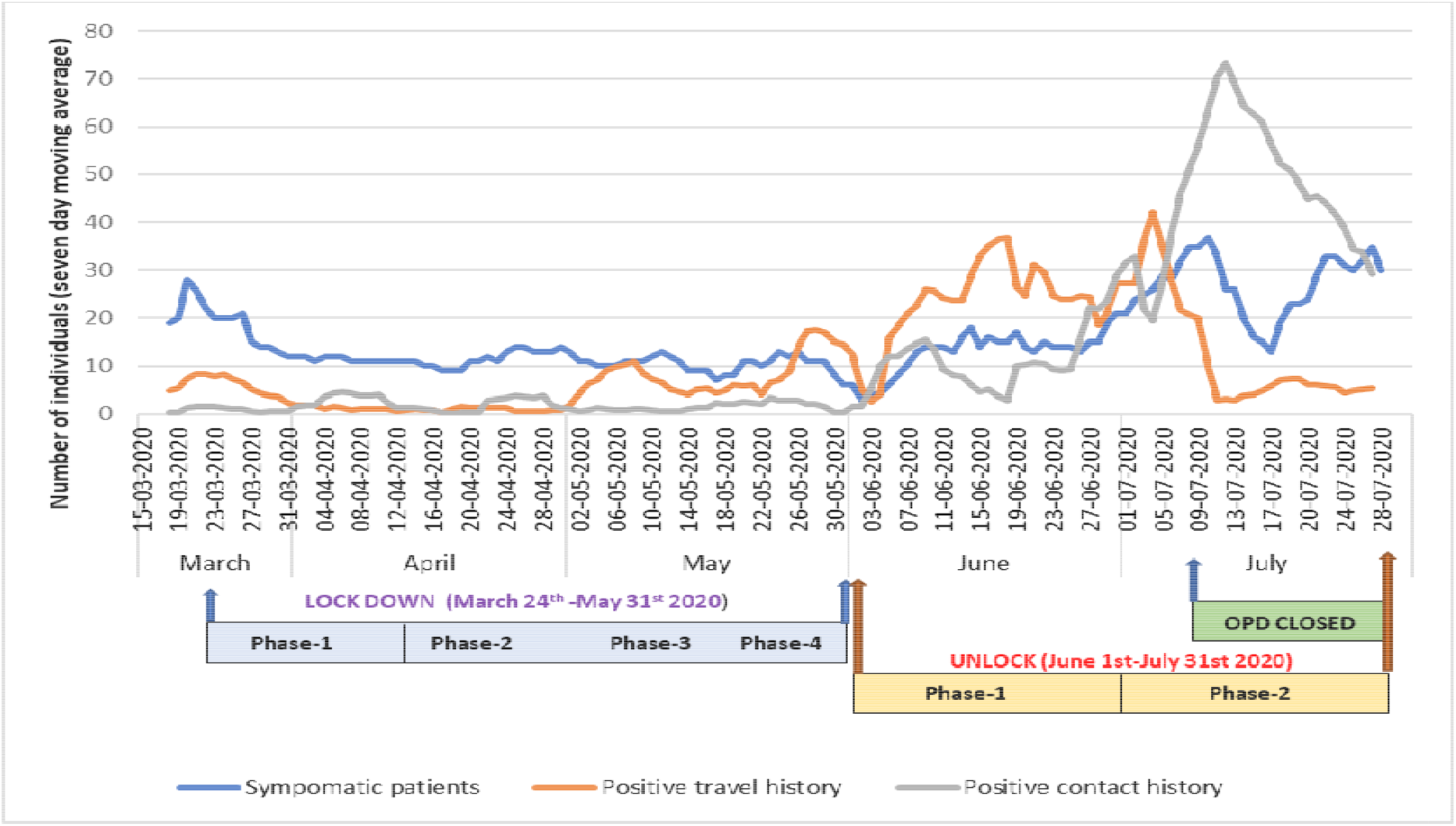
Time distribution of seven-day-moving average for number of individuals with positive travel history, positive contact history and symptoms:

The total daily patient visit showed a steady increase from April 2020 to 9^th^ July 2020 (range:7-268) followed by a steady decline till the end of July 2020 (range: 57-161). On the other hand, the weekly average daily sample testing increased significantly from June 2020 till mid of July 2020 (range:10.71-104.43) followed by a declining trend. Similarly, the seven-days moving average for positive cases (range:0-11.43) increased after from July 2020 onwards (range:3.57-11.43). **(Figure 5)**

**Figure 5:**
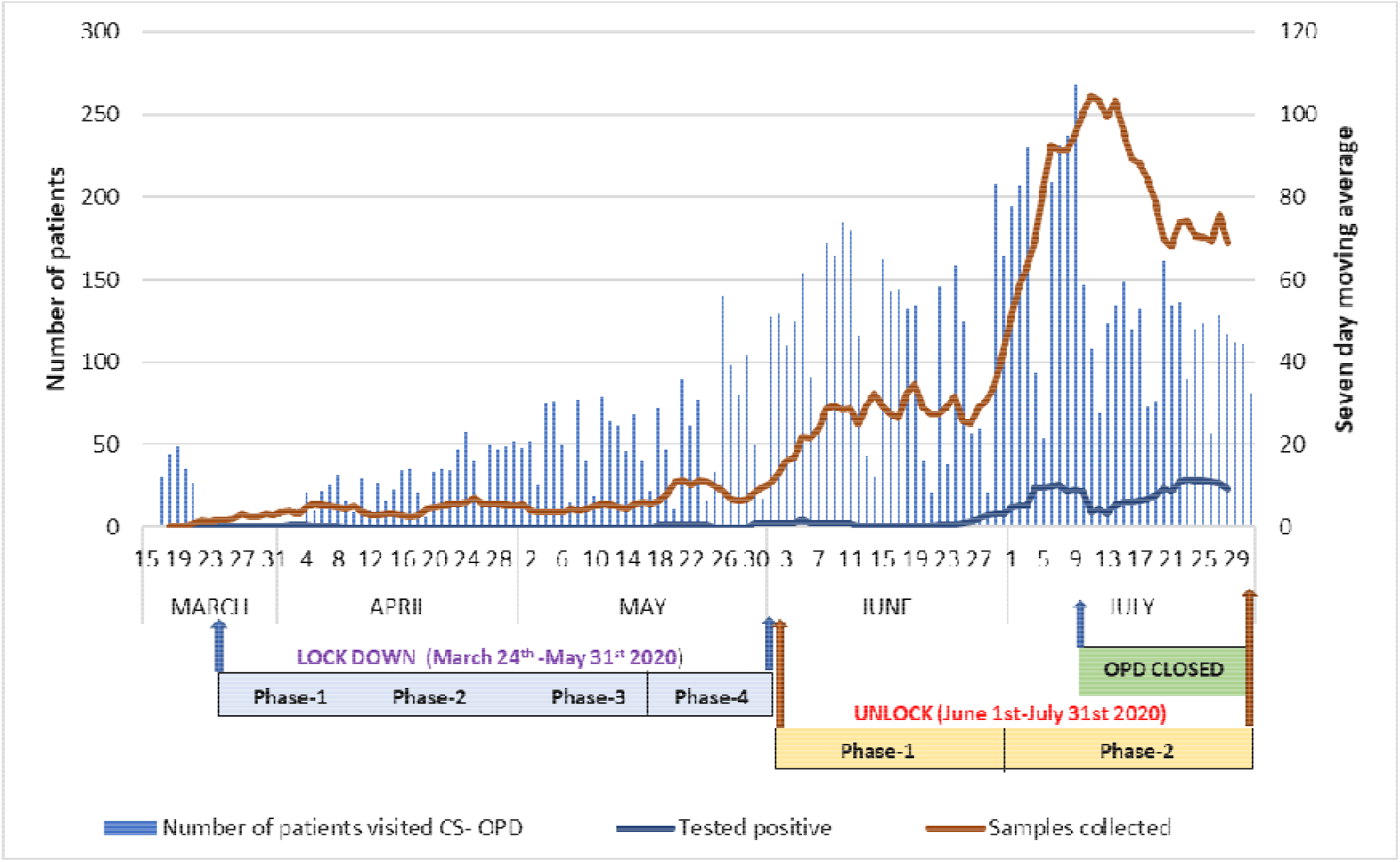
Variation of seven-day-moving average for number of samples tested and tested positive for COVID-19 as compared to daily patient visit.

**Table 2:**
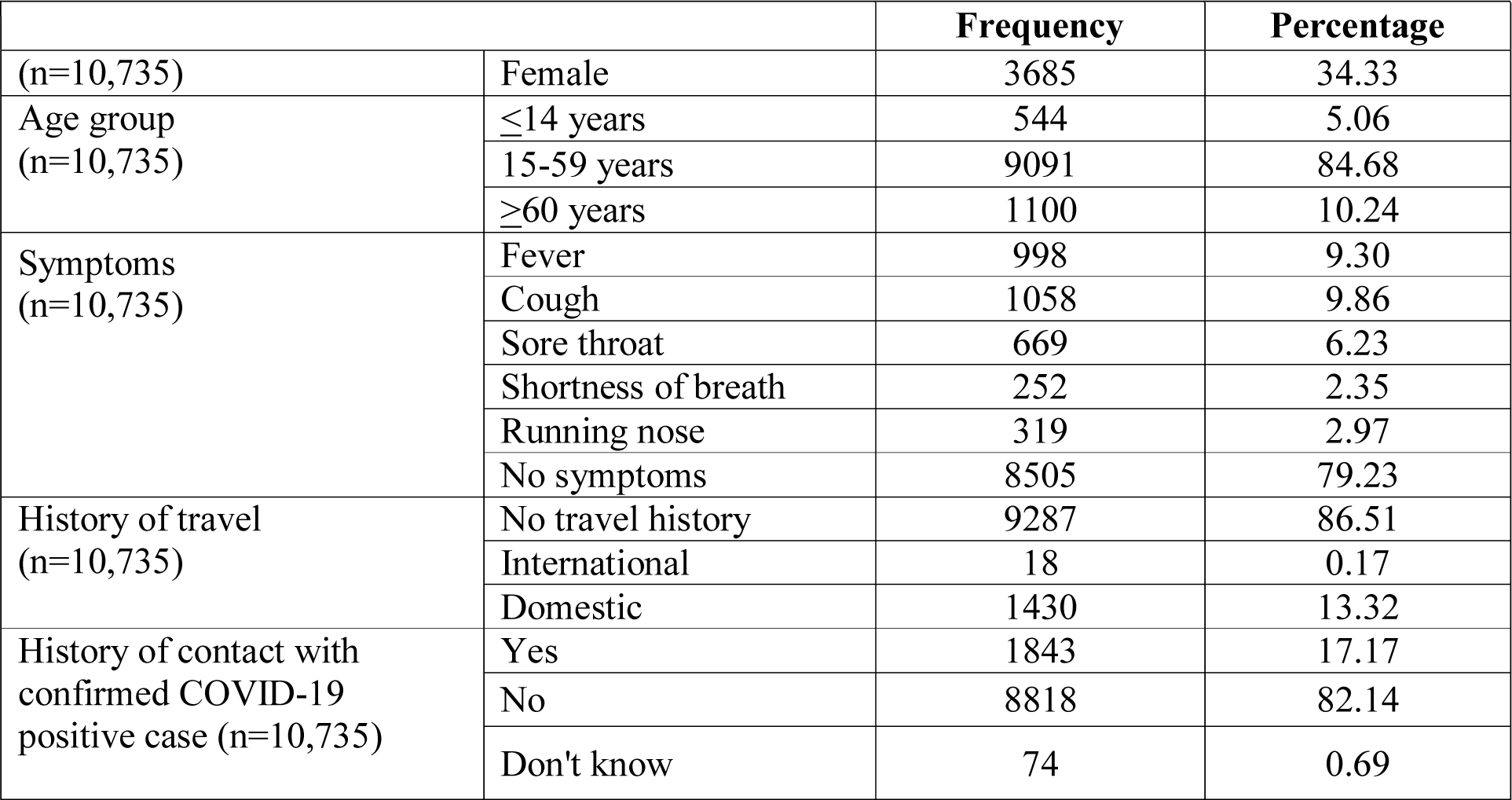
Distribution of CS-OPD patients based on symptoms, travel and contact history.

|  | Frequency | Percentage |
| --- | --- | --- |

|  |  |  |  |
| --- | --- | --- | --- |
| (n=10,735) | Female | 3685 | 34.33 |
| Age group<br>(n=10,735) | ≤14 years | 544 | 5.06 |
|  | 15-59 years | 9091 | 84.68 |
|  | ≥60 years | 1100 | 10.24 |
| Symptoms<br>(n=10,735) | Fever | 998 | 9.30 |
|  | Cough | 1058 | 9.86 |
|  | Sore throat | 669 | 6.23 |
|  | Shortness of breath | 252 | 2.35 |
|  | Running nose | 319 | 2.97 |
|  | No symptoms | 8505 | 79.23 |
| History of travel<br>(n=10,735) | No travel history | 9287 | 86.51 |
|  | International | 18 | 0.17 |
|  | Domestic | 1430 | 13.32 |
| History of contact with<br>confirmed COVID-19<br>positive case (n=10,735) | Yes | 1843 | 17.17 |
|  | No | 8818 | 82.14 |
|  | Don't know | 74 | 0.69 |

A total of 3652 samples were collected during the study period and a median of 10.00 (IQR=38.00) samples were being collected from CS-OPD every day. The overall testing rate and positivity rate for CS-OPD during this period were found to be 34.02% and 7.94% respectively. The patients from female paediatric population had the highest testing rate (44.51%) as well as positivity rate (18.18%). The overall testing and positivity rate was maximum for the paediatric age group. **(Table-3)**

**Table 3:** Age and gender-wise distribution of sample testing indicators indicators.

| Sl.No. | Age group | Gender | OPD visited<br>(Frequency) | Tested<br>(Frequency) | Tested<br>positive<br>(Frequency) | Testing<br>rate (%) | Positivit<br>y rate<br>(%) |
| --- | --- | --- | --- | --- | --- | --- | --- |
| 1 | <14<br>years | Male | 371 | 121 | 11 | 32.61 | 9.09 |
|  |  | Female | 173 | 77 | 14 | 44.51 | 18.18 |
|  |  | Total | 544 | 198 | 25 | 36.40 | 12.63 |
| 2 | 15-59<br>years | Male | 5898 | 1860 | 154 | 31.54 | 8.28 |
|  |  | Female | 3193 | 1303 | 86 | 40.81 | 6.60 |
|  |  | Total | 9091 | 3163 | 240 | 34.79 | 7.59 |
| 3 | >60<br>years | Male | 781 | 212 | 15 | 27.14 | 7.08 |
|  |  | Female | 319 | 79 | 10 | 24.76 | 12.66 |
|  |  | Total | 1100 | 291 | 25 | 26.45 | 8.59 |
| 4 | Total | Male | 7050 | 2193 | 180 | 31.11 | 8.21 |
|  |  | Female | 3685 | 1459 | 110 | 39.59 | 7.54 |
|  |  | Total | 10735 | 3652 | 290 | 34.02 | 7.94 |

The maximum proportion of samples were collected from Others category (29.71%) followed by Symptomatic healthcare workers Category (20.21%). However, the positivity rate was maximum in Symptomatic contacts of laboratory-confirmed cases (19.40%) followed by asymptomatic direct and high-risk contacts of laboratory-confirmed case – family members (18.82%). **(Table-4)**

**Table 4:** Category wise distribution of samples collected along with positivity rate-.

| Sl.No. | Category | Tested positive | Total samples | Positivity |
| --- | --- | --- | --- | --- |

|  |  | (% of total) | collected<br>(% of total) | rate(%) |
| --- | --- | --- | --- | --- |
| 1 | Symptomatic international travellers in the last 14 days | 2(0.68) | 18(0.49) | 11.12 |
| 2 | Symptomatic contacts of laboratory- confirmed case | 26(8.96) | 134(3.67) | 19.40 |
| 3 | Symptomatic healthcare workers | 37(12.75) | 738(20.21) | 5.01 |
| 4 | Patient with Severe acute respiratory infection | 3(1.03) | 54(1.48) | 5.56 |
| 5 | Asymptomatic high risk/direct contact of positive cases* | 0(0.0) | 42(1.15) | 0.00 |
| 6 | Asymptomatic direct and high-risk contacts of laboratory confirmed case – family members | 64(22.06) | 340(9.31) | 18.82 |
| 7 | Asymptomatic healthcare workers in contact with confirmed case without adequate protection | 28(9.65) | 732(20.04) | 3.83 |
| 8 | Symptomatic ILI in hospital | 16(5.51) | 147(4.03) | 10.88 |
| 9 | Pregnant women in/near labor | 2(0.68) | 30(0.82) | 6.67 |
| 9 | Symptomatic ILI among returnees and migrants within 7 days of illness | 11(3.79) | 88(2.41) | 12.50 |
| 10 | Symptomatic ILI in hotspot/containment zones | 23(7.93) | 233(6.38) | 9.87 |
| 11 | Others | 78(26.89) | 1085(29.71) | 7.19 |
| 12 | Not specified | 0(0.0) | 11(0.30) | 0.00 |
|  | **Total** | **290(100.00)** | **3652(100.00)** | **7.94** |

\* Asymptomatic high risk/direct contact of positive cases-This category was later divided in to same for healthcare workers (Category-5A) and family members (Category-5B) by ICMR.

## Discussion

The role of screening OPD for COVID -19 can be discussed from two perspectives, i.e. maintaining hospital function through preventing nosocomial infection and providing diagnostic services to suspected COVID-19 patients from the community in an accurate as well as timely manner. The previous experience during H1N1 pandemic as well as present experience with COVID-19 has reinforced the role of separate OPD under various names like “fever clinic”, “screening OPD”, “screening clinic” etc. (10)(11)(12) *Mahesh et al*. conducted a similar study in a tertiary care hospital of western India. They concluded that early diagnosis, quick initiation of treatment, infection control measures, and reasonable care at the hospital through flu OPD could effectively reduce morbidity and mortality during the H1N1 pandemic. (13) Similarly, in a study conducted by *Kwon et al*., they found that COVID-19 screening clinics were effective in maintaining the non-COVID-19 treatment facilities by reducing the incidence of nosocomial infection in the hospital. The CS-OPD might have played a crucial role in the prevention of possible nosocomial infection by early diagnosis and segregation of COVID-19 positive patients as well as health care workers at AIIMS Bhubaneswar. Although CS-OPD was planned and designed according to the existing health facility infrastructure and local environment, some of the improvements based on evidence from other studies can be incorporated in its functioning. Modifications like a separate passage for patients-staffs-wastes and sample collection in a negative pressure chamber can further strengthen the infection prevention and control measures. (10)(14)

As far as the CS-OPD patient profile is concerned, the maximum proportion of patient belongs to the male gender and are from 15-60 years age group. This is in line with the study done by *Khan et al*., and this may be due to lesser tendency among the female and elderly population to seek proactive COVID-19 related care because of social as well as inadequate health-seeking behaviour issues. (15)(11) Fever and cough were the two most common presenting symptoms among patients visiting CS-OPD which is similar to the findings of *Mohan et al*. in a study done among COVID-19 positive patients at a tertiary care institution in North India. (16) Maximum proportion (79.23%) of patients visiting CS-OPD was without any symptoms relevant to COVID-19. This asymptomatic population visiting CS-OPD was mainly consisted of patients coming to the hospital for follow up visits, patient attendants, healthcare workers and individual with travel history to hotspot areas at that point of time.

On further analysis of the profile of CS-OPD patients with respect to time, the number of patients visiting CS-OPD with travel history and contact history increased from June 2020. This may be due to the initiation of phased unlock after a nationwide lockdown from 25^th^ March 2020 to 31^st^ May 2020. (17) However, it again showed a declining trend from mid-July 2020. This can be attributed to the closure of AIIMS Bhubaneswar main OPD from 10^th^July 2020 due to staff crunch associated with increased COVID-19 infection among health care workers. (18) A similar type of trend was also observed with total samples collected as with the closure of OPD, the number of patients coming to AIIMS Bhubaneswar decreased suddenly leading to reduced screening and subsequent reduction in suspected cases. But the number of positive cases detected remained on the higher side despite OPD closure, mostly due to increasing case burden in the community.

In the case of testing indicators, the overall 7.94% positivity rate indicates that we may need a more efficient implementation of relevant public health interventions to reach the WHO devised epidemiological criteria for controlling the epidemic. (19) Higher positivity rate among the paediatric population is in contrast to findings from ICMR COVID study group where they reported a higher positivity rate among adult and elderly population. This may be due to higher testing rate among them as compared to other age groups. (20)

Apart from this, the ongoing evolution of ICMR testing strategy has also an impact on testing indicators as it has varied throughout the study period **(Figure-3)**. In spite of this, if we look into sampling categories, maximum samples have been contributed by the “others” category (29.71%). This may be explained by the fact that, at AIIMS Bhubaneswar being a tertiary care referral institution, patients with immunocompromised status like cancer (on chemotherapy/radiotherapy) or chronic kidney diseases (on dialysis) or patients with pre-operative conditions have been screened and tested extensively before admitting them to non-COVID wards. We included these patients under the “others” category as they didn’t fit to any other ICMR testing category. Similarly, the higher testing percentage among healthcare workers can be due to their increased susceptibility to infection, easy approachability to COVID-19 testing services and regular contact tracing at the institutional level.

On the other hand, maximum positivity among symptomatic contacts of laboratory-confirmed cases (19.40%) is in agreement with the ICMR COVID study group findings (10.6%). However, the second-highest positivity was detected in “Asymptomatic direct and high-risk contacts of laboratory-confirmed case – family members” (18.82%) in contrast to SARI patients (6.1%) in the ICMR COVID study group findings. This may be due to the fact that most of the SARI cases coming to the hospital were being referred directly to the emergency department for immediate care and proper evaluation. (20)

### Strengths and limitations

This is the one of the few studies from India which has been undertaken to understand the patient profile and functioning of a COVID-19 screening OPD in a tertiary healthcare setting. We have collected and analysed the data from all the patients who visited the screening OPD during the study period. Our study has been able to demonstrate the variation in patient profile associated with introduction of containment strategy to control COVID-19 pandemic. However, the periodic changes in testing strategy and categorisation of suspected cases for sample collection by ICMR had influenced the testing and positivity rate in our study.

## Conclusion and Recommendations

Our study gives an overview of the functioning of a COVID-19 screening OPD as a part of a tertiary care institution. We have demonstrated how public health intervention like lockdown and travel restrictions impacted the patient profile and brought a change in its due course of the study period. Although the COVID-19 screening OPD has been effective in providing screening and diagnostic services to patients, various best practices, and evolving strategies based on evidence should be added to it continuously. Inclusion of point-of-care testing services and broadening the ambit of suspected criteria at screening OPDs can help us to detect more COVID-19 positive cases. Moreover, tertiary care institutions should also plan permanent and separate infectious disease clinics like CS-OPDs and keep on upgrading them to address evolving public health challenges like COVID-19.(21) As the pandemic continues, it is evident that no single strategy is sufficiently effective. Therefore, the health system must adhere to a holistic approach in dealing with this pandemic, for which COVID-19 screening OPDs remains a critical component.

## Data Availability

All the data is with the corresponding author and co-author.

## Declaration of conflict of interest

Authors declare that they have no conflicts of interest to disclose.

### Acknowledgements

We sincerely thank to all the participants for their participation. We are thankful to all the staffs of COVID-19 screening OPD of All India Institute of Medical Sciences, Bhubaneswar for their co-operation to conduct the study.

## Authors’ contribution

SKP: Contributed in conception and designing, analysis and interpretation of data and design of the study and drafting of article.

DPS1: Contributed in data acquisition, revising the manuscript for important intellectual content.

DPS2: Contributed in data acquisition, revising the manuscript for important intellectual content.

AKS: Contributed in conception and designing of the study, analysis and interpretation of data and revising the manuscript for important intellectual content.

BKP: Contributed in conception and designing of the study, analysis and interpretation of data and revising the manuscript for important intellectual content.

SM: Contributed in conception and designing of the study, facilitating data collection and revising the manuscript for important intellectual content.

All the authors gave final approval of submitted version.

